# Longitudinal Analysis of the Determinants of Life Expectancy and Healthy Life Expectancy: A Causal Approach

**DOI:** 10.1101/2021.09.09.21263169

**Authors:** Rohan Aanegola, Shinpei Nakamura-Sakai, Navin Kumar

## Abstract

Understanding the determinants of health is essential to designing effective strategies to advance economic growth, reduce disease and disability, and enhance quality of life. We undertake a comprehensive outlook on public health by incorporating three metrics - life expectancy (LE), healthy life expectancy (HLE), and the discrepancy between the two. We investigate the effects of various health and socio-economic factors on these metrics and employ causal machine learning and statistical methods such as propensity score matching, X-learners, and causal forests to calculate treatment effects. An increase in basic water services and public health expenditure significantly increased average LE, whereas high HIV prevalence rates and poverty rates reduced average LE. High GNI per capita and moderate BMI increased HLE while high HIV prevalence rates decreased HLE. High public health expenditure and high GNI per capita expand the gap between HLE and LE, whereas high HIV prevalence rates and moderate BMI diminish this gap. Results suggest that policymakers should utilize governmental resources to improve public health infrastructure rather than provide fiscal incentives to encourage private healthcare infrastructure. Additionally, more emphasis should be put on increasing educational levels of the general public by increasing educational expenditure and making educational institutions, public and private, more accountable.

## 1 Introduction

Life Expectancy (LE) at birth is one of the key metrics for assessing population health and hence, of primary interest in medical research [1]. A study by the WHO reported that people are healthier, wealthier, and living longer today than 30 years ago [2]. The high increase in LE is generally attributed to improvements in access to clean water, medical and technological advancements, eradication of life-threatening diseases, and increases in agricultural development. The advancement of human health is considered as one of the key policies for sustainable development [3]. There is also a growing consensus that improving health could advance economic growth [4]. A report by the WHO in 2001 stated that extending health services to the world’s poor could save millions of lives each year, reduce poverty, spur economic development, and promote global security. Health outcomes also affect human capital investment, inter-generational transfers, and incentives for pension benefit claims [5].

The primary focus of prior research has been the identification of correlations between different indicators of LE and the metric itself. Kaplan et al. qualitatively revealed that higher educational attainment correlates to higher life expectancies [6]. A study by Khang et al. revealed that there was a positive impact on LE from increased incomes [7]. Healthcare services such as an increased number of physicians, hospital beds, and prenatal examinations could reduce mortality and increase LE [8, 9]. Mondal et al. suggested that HIV prevalence rate, mean years of schooling, and GNI capita are significant predictors of LE in the low and lower-middle-income countries [5]. Martin et al. showed that per capita income, rate of hospital beds, medical staff, and nurses Granger-cause the variable ‘life expectancy at birth’ [10]. A negative association was found between unemployment and health outcomes [11].

While LE captures the mortality of a population, it fails to account for morbidity - there exist several diseases that aren’t potentially life-threatening, but can severely hinder lifestyle. This limitation is overcome by Healthy Life Expectancy (HLE), defined as the average number of years that a person can expect to live in full health not hampered by disabling illness and injuries. HLE is a superior measure to LE as it combines length and quality of life into a single measure [12]. Medical advancements and improvements in standards of living have meant that people are living longer, but the 2015 Global Burden of Disease data showed that the corresponding increase in HLE was significantly less, meaning that people have longer but less healthy lives [13].

Although evidence of the effects of healthcare resources, economic development, and educational attainment on LE has been proved in previous studies [5, 6, 10], there is limited research on the determinants of HLE and causes of the discrepancy between LE and HLE. Furthermore, the main focus of existing literature has been on the identification of correlations between health indicators and LE. While this may provide useful insight, the results are often misleading. Due to the complex nature of the current health context, there is significant confounding between variables of different types, making correlation analyses to attest each variable’s effect in isolation unreliable. Our current study endeavors to bridge this gap in literature by presenting a causal analysis of the effects of various health and socio-economic factors on both, LE and HLE. In addition to this, we determine covariates that have a direct causal effect on the discrepancy between the two measures (section 3). To gauge the relative impact of each covariate, we make use of 3 different causal machine learning/ statistical models stated in increasing order of complexity: propensity score matching, X-learners, and causal forests (section 3). By implementing three different models and achieving analogous results, we validate the robustness of our results.

## 2 Data

For the analysis, we used WHO and UNSECO databases (*n* = 3111) consisting of 30 continuous covariates and the response variable, LE at birth, recorded by 183 countries during the years 2000-2016. Covariates largely consisted of quantitative health and socio-economic factors (see section A of appendix). Data of HLE of 30 European countries during the years 2008-2019 were acquired using a database available at Eurostat (*n* = 270).

## 3 Methods

Missing values were imputed using the Multiple Imputation by Chained Equations algorithm (MICE) [14]. After observing the distributions of all covariates to be moderately Gaussian, we concluded that the data were missing at random and set the minimum and maximum prediction values to be the minimum and maximum existing values of covariates in the dataset, respectively [15]. We applied *log* transformations on some covariates to decrease the variability and make them conform more closely to the normal distribution [16]. All covariates were adjusted to have a mean of 0 and unit standard deviation to make results more interpretable and to reduce errors in regression models. To minimize multicollinearity as well as deal with high dimensionality, we used L1 regularized (lasso) regression wherein the weight matrix is calculated as

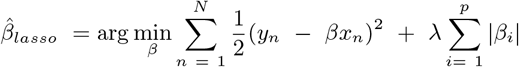

where *N* is the number of samples, *p* is the number of features, *y* is the ground truth, and *x* is the feature matrix. The penalty term *λ* regularizes the magnitude of parameter values, shrinking some parameters to zero. Using cross validated grid search, we found the most optimal value of *λ* to be 0.1 and arrived at weakly correlated covariates. All selected features were made to act as treatments individually while keeping the rest as covariates. Treatments were then binarized in order to mitigate the effects of significant outliers and to make results more comprehensive.

Bayesian Networks were generated using Microsoft’s DoWhy [17] to check for confounding and to understand conditional dependencies between covariates, the treatment, and the outcome. Causal effects were estimated using three treatment effects - ATE (Average Treatment Effect), ATT (Average Treatment Effect on the Treated), and ATC (Average Treatment Effect on the Control) - defined as follows:

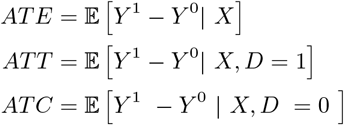

where D denotes treatment indicator, *Y* ^1^ denotes outcome with treatment, and *Y* ^0^ denotes outcome without treatment.

Three methods were used to test the causal impact of all the treatments selected, starting from the least sophisticated (propensity score matching) to the most sophisticated (causal forest).

### 3.1 Propensity Score Matching

As data are from observational studies, there is no random assignment, making it almost impossible to eliminate confounding. The propensity score, defined by Rosenbaum and Rubin (1983) [18], is the probability of treatment assignment conditional on observed baseline covariates or, equivalently, ℙ (*D* = 1|*X*). It is generally calculated using a logistic function

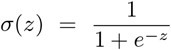

Rather than controlling for each covariate, controlling for the propensity score alone is sufficient to avoid confounding and bias. To calculate propensity scores, we first used logistic regression setting the outcome as the treatment (dichotomous) and the other covariates as arguments. However, due to the linearly separable nature of all the covariates, we obtained perfect quasi separation, making scores difficult to match. Therefore, we made use of the bias reduction method of Firth [19], a procedure that returns estimates with improved bias and MSE that are always finite even in cases where the maximum likelihood estimates are infinite (quasi separation).

We formed one-to-one paired matched sets of treated and untreated subjects with similar propensity scores. Treatment effects were calculated by comparing outcomes between treated and untreated subjects in the matched sample. Our analysis of the propensity score matched sample mimics that of a randomized control trial as one can directly compare outcomes between treated and untreated subjects within the propensity score matched sample [20].

### 3.2 X-Learner

The X-learner, first introduced by Kunzel et al [21], is a meta-learner used to estimate heterogeneous treatment effects (see section B.1 of appendix) The CATE was estimated using the following algorithm:

1. Estimate the average outcomes:

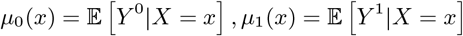
2. Compute imputed treatment effects:

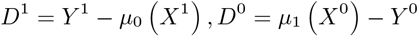
3. Estimate CATE in two ways:

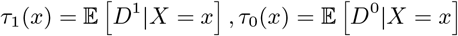
4. Take weighted average of the estimates:

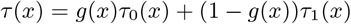

*g*(*x*) is a weighting function chosen to minimize *τ* (*x*) for which we made use of the propensity score. From the CATE estimated, the ATE, ATT, and ATC were computed. We used multiple linear regression to estimate the conditional expectations of the outcomes separately for the treatment and control groups.

### 3.3 Causal Forests

Athey et al [22] modified Conditional and Regression Trees (CARTs) into causal trees and combined causal trees into causal forests to produce estimates that can be used for valid statistical inference.

We implemented honest causal forests wherein subsamples were randomly split in half – the first half was used when performing splitting while the second half was used to populate the tree’s leaf nodes. By using different subsamples for constructing the tree in the causal forest and for making predictions, we made our model less prone to overfitting [23]. Applying this novel empirical methodology, we were able to examine the causal effects of each treatment on the three outcome variables.

## 4 Results

Estimates of ATE, ATT, and ATC along with their means were calculated for each response variable: LE, HLE, and LE – HLE. Summaries of the different treatment effects (ATE, ATC, ATT) for all 3 response variables are summarized in tables 1-3. For significance tests, refer to section C of appendix.

**Table 1:** LE

| Treatment | Prop-Score |  |  | X-Learner |  |  | Causal Forest |  |  | Mean |  |  |
| --- | --- | --- | --- | --- | --- | --- | --- | --- | --- | --- | --- | --- |
|  | ATE | ATC | ATT | ATE | ATC | ATT | ATE | ATC | ATT | ATE | ATC | ATT |
| BWS | 4.29 | 6.63 | 1.94 | 4.24 | 4.20 | 1.37 | 3.35 | 3.07 | 2.66 | 3.96 | 4.63 | 1.99 |
| GNIC | 2.02 | 0.76 | 3.29 | 2.36 | 1.53 | 3.17 | 1.72 | 0.47 | 2.91 | 2.04 | 0.92 | 3.12 |
| HIVPR | -3.04 | -2.42 | -3.66 | -2.81 | -2.01 | -3.61 | -3.88 | -3.83 | -3.79 | -3.24 | -2.75 | -3.69 |
| LIT | 1.95 | 3.15 | 0.74 | 1.81 | 2.76 | 1.86 | 1.08 | 2.09 | 0.60 | 1.61 | 2.67 | 1.07 |
| POV | -2.22 | -0.36 | -4.08 | -1.94 | -0.89 | -3.99 | -1.36 | 0.25 | -2.98 | -1.84 | -0.33 | -3.68 |
| PUBHE | 1.24 | 0.83 | 3.31 | 1.60 | 1.40 | 1.79 | 1.88 | 1.67 | 2.08 | 1.57 | 1.30 | 2.39 |

**Table 2:** HLE

| Treatment | Prop-Score |  |  | X-Learner |  |  | Causal Forest |  |  | Mean |  |  |
| --- | --- | --- | --- | --- | --- | --- | --- | --- | --- | --- | --- | --- |
|  | ATE | ATC | ATT | ATE | ATC | ATT | ATE | ATC | ATT | ATE | ATC | ATT |
| BMI | 2.98 | 3.54 | 2.31 | 2.96 | 2.09 | 2.16 | 2.84 | 3.88 | 1.58 | 2.93 | 3.17 | 2.02 |
| EDUE | 1.44 | 1.73 | 1.15 | 2.63 | 2.74 | 2.57 | 1.52 | 2.14 | 1.89 | 1.86 | 2.20 | 1.87 |
| GNIC | 4.18 | 2.99 | 5.36 | 3.46 | 3.41 | 3.50 | 3.03 | 3.08 | 3.94 | 3.55 | 3.16 | 4.27 |
| HIVPR | -0.47 | 0.10 | -1.04 | -0.85 | -0.81 | -0.88 | -0.14 | -0.39 | -0.12 | -0.49 | -0.37 | -0.68 |
| PRIVHE | 1.21 | 4.51 | -2.08 | 1.55 | 2.61 | -1.50 | 1.42 | 2.47 | -1.30 | 1.39 | 3.20 | -1.63 |

**Table 3:** LE-HLE

| Treatment | Prop-Score |  |  | X-Learner |  |  | Causal Forest |  |  | Mean |  |  |
| --- | --- | --- | --- | --- | --- | --- | --- | --- | --- | --- | --- | --- |
|  | ATE | ATC | ATT | ATE | ATC | ATT | ATE | ATC | ATT | ATE | ATC | ATT |
| BMI | -2.56 | -3.06 | -1.94 | -2.62 | -3.58 | -1.79 | -2.09 | -2.56 | -1.51 | -2.42 | -3.07 | -1.75 |
| EDUE | -1.25 | -0.98 | -1.51 | -0.97 | -0.98 | -0.96 | -1.23 | -0.43 | -0.42 | -1.15 | -0.80 | -0.97 |
| GNIC | 1.92 | 1.13 | 2.72 | 0.53 | 0.71 | 0.33 | 0.55 | 0.55 | 0.55 | 1.00 | 0.79 | 1.20 |
| HIVPR | -0.02 | -0.62 | -0.58 | -0.98 | -1.26 | -0.68 | -0.01 | -0.36 | -0.35 | -0.34 | -0.74 | -0.53 |
| PRIHE | -0.82 | -2.63 | 1.22 | -1.02 | -1.83 | 0.27 | -0.55 | -1.95 | 0.16 | -0.80 | -2.14 | 0.55 |
| PUBHE | 1.34 | -0.22 | 2.89 | 1.00 | 0.20 | 1.85 | 1.26 | -0.28 | 1.79 | 1.20 | -0.10 | 2.18 |

**Table 4:** Body Mass Index Classification

| BMI | Category |
| --- | --- |
| < 18.5 | Underweight |
| 18.5 – 24.9 | Healthy Weight |
| 25 – 29.9 | Overweight |
| > 30 | Obese |

### Basic Water Services (BWS)

Access to basic water services is intrinsic to good health and thus, had the largest impact on LE. It had the highest ATE (3.96) and ATC (4.63), but a relatively lower ATT (1.99). This means that the population that had access to basic water services had an increased 1.99 years of LE as a result, whereas the population that did not have access to basic water services could have gained 4.63 extra years if they had access. However, it had a negligible impact on HLE and LE - HLE (feature was not selected by lasso).

### BMI (BMI)

The widely accepted ranges of BMIs are as follows:

BMI significantly impacted HLE (ATE = 2.93) and decreased the gap between LE and HLE (ATE = -2.42). Having achieved a median BMI of 20.4, we assumed (for the analysis) that countries’ populations that did not receive the treatment (BMI < *median* (BMI)) were underweight.

### HIV Prevalence Rate (HIVPR)

HIV PR had a significant impact on all 3 response variables. Out of the four diseases included in the analysis (Diphtheria, Measles, Polio, and HIV), HIV had the most negative causal impact on all three response variables. It, however, had a much smaller impact on HLE (ATE = -0.49) and LE – HLE (ATE = -0.34) than on LE alone (ATE = -3.24).

### Educational Expenditure (EDUE) and Literacy (LIT)

We took into account three educational variables into our analysis – Educational Expenditure, Mean years of schooling, and literacy rate. However, mean years of schooling was eliminated during feature selection. High literacy rates had a positive impact on LE (ATE = 1.61) but negligible effects on HLE and LE – HLE. Conversely, educational expenditure increased HLE (ATE = 1.86) and reduced LE-HLE (ATE = -1.15), but had an insignificant effect on LE.

### Public Health Expenditure (PUBHE), Private Health Expenditure (PRIHE), and GNI Capita (GNIC)

We took public and private health expenditures as two distinct variables rather than taking ‘total health expenditure’ to contrast the impacts of the private and public sectors. While public health expenditure did not affect HLE, it had a positive effect on LE (ATE = 1.57). However, the converse was true for private health expenditure – It showed a positive effect on HLE (ATE = 1.39) but not on LE. GNI per Capita, like HIV PR, had a significant impact on all 3 response variables. It had a much more significant impact on HLE (3.55) than on LE (2.04) and LE – HLE (1.00).

## 5 Discussion

In general, countries with basic water services, low poverty rates, and low HIV prevalence rates have higher LE, while countries with high GNI per capita, high educational expenditure, and BMI not considered underweight have high HLE. Moreover, BMI, GNI capita, and public health expenditure are the most significant factors that cause the disparity between LE and HLE.

Our results are largely consistent with prior research. Research on the effects of HIV on LE observed a 0.34-year decrease in LE due to HIV PR in low and middle-income countries [24]. Hassan et al found that patients with the HIV infection suffered a 36-year loss in LE [25]. Our results on literacy and educational expenditure are in accordance with past research - Luy et al. [26] demonstrated that higher educational levels increased LE by 2.8 years in the US during the years 1990-2010. Kaplan et al. argued that while cancer screening and optimizing established risk factors for premature death typically extent LE by less than a year, remediating the health disparity associated with low educational attainment might enhance LE by up to a decade [6]. Previous work on the effects of public health expenditure and private health expenditure, however, has been conflicted. For example, Barenberg et al.[27] found inverse results of the relationship between public health spending and infant mortality rate for 176 countries. Furthermore, analyses by Novignon et al. in sub-Saharan Africa [28] and Ethat et al. in eastern Mediterranean countries [29] concluded that public health care spending had a relatively higher impact than private health spending on health status. However, Raeesi et al suggested that the effect of private health expenditures on health outcomes in countries was higher than public health expenditures [30].

A particularly unanticipated result obtained was during feature selection for LE. It was observed that public health expenditure had a stronger correlation to LE (0.60) than private health expenditure (0.28), and additionally, lasso selected Public HE and disregarded Private HE. This empirical observation, grounded by results of the causal analysis (Public HE ATE =1.237), has profound implications – The expenditure by the government on health infrastructure and resources proves to be more impactful than the expenditure by any other external body, even at the individual level, contributing to the economy, on LE. However, the converse is true for HLE – private HE was observed to be more significant than public HE. Both public HE and private HE are strong explanatory variables of the discrepancy between LE and HLE. The positive sign of ATE for public HE is because it has a strong effect on LE and a minimal effect on HLE, while the converse is true for private HE.

One possible explanation for our results is that private HE is over-represented by the expenditure of the high-income population and very minimally, that of the lower-income population, leading to a skewed and unrepresentative measure of health infrastructure. Therefore, we suggest that governing bodies utilize more of their own resources to improve public health infrastructure rather than provide fiscal incentives to encourage private healthcare infrastructure. This could be since government spending is generally both inefficient and inequitable [31]. While this seemingly does not impair treatment of fatal diseases that lead to mortality, it could hinder treatment of non-fatal diseases that have significant effects on lifestyles (high blood pressure, diabetes, scoliosis, etc.), reducing HLE.

The high negative impact of high HIV prevalence rates on LE and HLE is due to the virus’s inherent potency. It is well established that HIV is a deadly viral disease that eventually attacks the immune system of the infected individual. The median survival time with HIV (untreated) is 8-10 years [32], drastically reducing LE. The virus’s fatality is further exacerbated by the absence of an effective cure – there exist only methods to control it. Furthermore, Most people living with HIV struggle to attend to daily tasks of living and participate in moderate to vigorous physical activities [33] resulting in shortened HLE.

A probable explanation for the positive effect of educational expenditure and literacy rates on LE and HLE is that people with more education tend to be more conscious of their health and can comprehend information related to nutrition and hygiene better. It enhances a sense of personal control that encourages a healthy lifestyle with ample physical exercise and a healthy diet. Also, higher educational levels are typically associated with timely receipt of healthcare, reducing fatality and risk of chronic conditions. Thus, due to the strong associations between education and population health, educational policies could also serve as indirect health policies

GNI per capita is the most prevalent measure of economic development of a country [34]. It is a major determinant of LE and HLE due to its strong association with greater advancements in medical technologies institutional improvements, improvements in living conditions (housing, health services, etc.), and lower mortality rates. People with higher wages would be able to afford better medical treatment, have access to good medical infrastructure and receive timely healthcare. Another explanation behind the connection between GNI per capita and LE is the effect of food supply on mortality. There have been statistically significant parallels drawn between prices of food and mortality [35]. This is also a viable explanation for the strong impact on HLE – lack of access to food could lead to malnourishment and other maladies inhibiting lifestyle. An effective solution is to increase health care expenditure itself since this not only increases LE and HLE directly but also indirectly by increasing GNI, as evidenced by [36].

Our results on the effect of BMI on LE and HLE are backed by a study performed by Ochiai et al. [37] which explained that underweight status was associated with scoliosis and osteoporosis – diseases that are not potentially life-threatening but can severely affect standards of living (affecting HLE and not LE). Governments should recognize that good nutrition is a priority for national health and prioritize nutritional quality in food assistance programs.

## 6 Conclusion

This study explored the causal impact of several health and socio-economic factors on three outcomes: Life Expectancy, Healthy Life Expectancy, and the gap between life expectancy and healthy life expectancy. We undertake a comprehensive outlook on public health to present an exhaustive analysis and highlight the direction of health policy in the current world.

Results suggest that an increase in basic water services and government health expenditure significantly increase average LE, whereas high HIV prevalence rates and poverty rates reduce average LE. High GNI per capita and moderate BMI increase HLE while high HIV prevalence rates decrease HLE. high public health expenditure and high GNI per capita expand the gap between HLE and LE (meaning that they affect the outcomes differently), whereas high HIV prevalence rates and moderate BMI diminish this gap. Overall, our research offers several important findings that could enhance policymakers’ and local authoritative bodies’ understanding of which factors influence public health the most and mandate accordingly.

The main restriction of this paper lies in the limited number of observations available for HLE, consequently restricting our analysis of this metric to only 30 European countries. Another drawback is that we did not differentiate between unhealthy BMIs > 25 and healthy BMIs between 20.4 and 24.9 and treated them equivalently. Also, a person can have a low BMI as a result of healthy living and not just malnutrition or disease [38].

Finally, future research should incorporate other factors that may affect LE and HLE such as climate, happiness, and demographics. Further, gender stratification could be performed to assess LE and HLE separately for males and females to identify disparities if any.

## Data Availability

Data are from WHO, Kaggle, and Eurostat databases.

## Acknowledgements

We received no funding sources. We thank the editor and reviewers for their comments.

## A Definitions

- Literacy Rate: The percentage of population aged 15 years and over who can both read and write with understanding a short simple statement on their everyday life. Generally, literacy also encompasses ‘numeracy’, the ability to make simple arithmetic calculations.
- HIV Prevalence Rate: The estimated number of adults aged 15-49 years with HIV infection, whether or not they have developed symptoms of AIDS, expressed as a percentage of the total population in that age group.
- Private Health Expenditure: The level of general government expenditure on health (GGHE) expressed as a percentage of total government expenditure.
- Public Health Expenditure: The expenditure on health care incurred by public funds.
- Poverty: The proportion of employed population below $1.90 purchasing power parity (PPP).
- Basic Water Services as a Percentage: The percentage of population using at least basic drinking water services. Basic drinking water services is defined as drinking water from an improved source, provided collection time is not more than 30 minutes for a round trip.
- GNI per capita: The gross national income, converted to U.S. dollars using the World Bank Atlas method, divided by the midyear population. GNI is the sum of value added by all resident producers plus any product taxes (less subsidies) not included in the valuation of output plus net receipts of primary income (compensation of employees and property income) from abroad.
- Educational Expenditure: The amount of total government’s budget of a country allotted to education.
- BMI: It is a measure of nutritional status in adults calculated by dividing a person’s weight in kilograms by the square of the person’s height in meters (kg/m2)

## B Model Descriptions

### B.1 X-Learner

The X-Learner, introduced by Kunzel et al, is a metalearner used for estimating Conditional Average Treatment Effect in a binary treatment setting. Metalearners decompose estimating the CATE into several sub-regression problems that can be solved with any regression or supervised learning method (Examples include the X-Learner, T-Learner, R-Learner, and S-Learner.). The X-Learner, built upon the T-Learner, is especially powerful since it can translate any supervised learning or regression algorithm into a CATE estimator. The X Learner has two advantages over the estimators of the CATE. First, it can adapt to structural properties such as the smoothness of the CATE (which is useful since the CATE is often zero or approximately linear). Second, it is effective when the number of units in the treatment groups is different. Implementation of the X-learner is done using the algorithm mentioned in section 3.

### B.2 Causal Forest

The causal forest uses the framework of the generalized random forest introduced by Athey et al. [22] to provide valid statistical inference. This model builds upon several properties of the random forest introduced by Breiman [39] such as random split selection and sub-sampling. While a random forest is composed of decision trees, a causal forest is an ensemble of causal trees, where each causal tree learns a low-dimensional representation of treatment effect heterogeneity., The objective of the split criterion within each tree is to capture heterogeneity in the parameter of interest rather than to minimize the predictive error [40]. A key characteristic of the causal forest is that the forest predictors are not computed by averaging treatment estimates, but rather by the nearest neighbor mechanism. To deal with overfitting, causal forests use an honesty condition - sample splitting is used to create honest trees where half the data is used to estimate the tree structure (splitting subsample) and the other half is used to estimate the treatment effect in each leaf (estimating subsample). Thus, by using different subsamples for constructing the tree in the causal forest and for making predictions, the model reduces overfitting.

## C P-Values

We define the null and alternate hypotheses as follows:

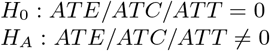

Tables 5-7 show the result of the hypothesis test (tested at the 10% significance level). The null hypothesis is rejected for most of the variables, therefore supporting our argument.

**Table 5:** LE

| Treatment | p-value |  |  |
| --- | --- | --- | --- |
|  | ATE | ATC | ATT |
| BWS | 0.00 | 0.00 | 0.12* |
| GNIC | 0.00 | 0.15* | 0.00 |
| HIVPR | 0.00 | 0.00 | 0.00 |
| LIT | 0.00 | 0.00 | 0.43* |
| POV | 0.03 | 0.65* | 0.03 |
| PUBHE | 0.00 | 0.21* | 0.00 |

**Table 6:** HLE

| Treatment | p-value |  |  |
| --- | --- | --- | --- |
|  | ATE | ATC | ATT |
| BMI | 0.00 | 0.00 | 0.00 |
| EDUE | 0.00 | 0.00 | 0.02 |
| GNIC | 0.04 | 0.08 | 0.14* |
| HIVPR | 0.14* | 0.16* | 0.02 |
| PRIVHE | 0.06 | 0.09 | 0.04 |

**Table 7:** LE-HLE

| Treatment | p-value |  |  |
| --- | --- | --- | --- |
|  | ATE | ATC | ATT |
| BMI | 0.00 | 0.00 | 0.07 |
| EDUE | 0.13* | 0.31* | 0.13* |
| GNIC | 0.17* | 0.55* | 0.14* |
| HIVPR | 0.09 | 0.34* | 0.02 |
| PRIHE | 0.07 | 0.15* | 0.07 |
| PUBHE | 0.09 | 0.12* | 0.05 |

**(*) means that for the corresponding variables, the null hypothesis is not rejected**.

